# Hypertension and Dementia: A Mendelian Randomisation Study Assessing Potential Causal Relationships with All-Cause Dementia, Alzheimer’s Disease and Vascular Dementia in the UK Biobank

**DOI:** 10.1101/2025.03.12.25323830

**Authors:** Roopal Desai, Joel Heller, Rin Wada, Michael Anis Mihdi Afnan, Charlene Cheong, Sijun Zhang, Fulvio Deo, Michelle Tam, Emma Anderson, Joshua Stott, Georgina Charlesworth, James W Yarmolinsky, Paul Elliott, Marc Chadeau-Hyam, Verena Zuber

## Abstract

Hypertension is a key modifiable risk factor for longevity. However, evidence linking hypertension to dementia remains inconsistent, potentially due to subtype-specific effects. We aimed to examine relationships between systolic blood pressure (SBP) and risk of Alzheimer’s disease (AD) and vascular dementia (VaD) using individual-level Mendelian randomization (MR) analyses. Logistic and Cox regressions assessed the association of blood pressure traits with all-cause dementia, AD and VaD in the UK Biobank cohort. Observational analyses indicated SBP was associated with increased risk of all dementia outcomes. MR analyses indicated genetically proxied SBP was associated with increased risk of all-cause dementia (OR=1.26, [95% CI:1.15;1.38]; HR=1.28, [95% CI:1.17;1.40]). When stratified by subtype, higher SBP was associated with increased risk of VaD (OR=1.56, [95% CI:1.25;1.93]; HR=1.58, [95% CI:1.28;1.97]), but not AD (OR=1.10, [95% CI:0.95;1.26]; HR=1.11, [95% CI:0.97;1.28]). Post-hoc analyses supported a differential effect. Hypertension appears to be a greater risk factor for VaD than AD.

## 1. Introduction

Dementia is a rapidly escalating global health challenge, affecting over 50 million people worldwide, with projections suggesting a tripling of prevalence by 2050^1^. To date, the advancement of pharmaceutical interventions has been slow with the current first generation of drugs slowing, without halting or reversing, the decline in Alzheimer’s disease (AD). Therefore, prevention remains a key global health priority. Hypertension is one putative modifiable risk factor that has received much attention. Mid-life hypertension is reported to be one of the 14 most important modifiable lifestyle dementia risk factors, increasing risk of dementia and accounting for up to 2% of incident dementia cases^2^. The evidence for the link between hypertension and dementia comes from a range of different approaches including observational studies, randomised controlled trials (RCT) and Mendelian randomization (MR) studies. However, the evidence from different study types has yielded conflicting results. Resolution of this issue is very important to understand the public health impact on dementia risk of blood pressure control measures, as currently guidelines are mostly based on observational studies from which we cannot infer causality.

Observational studies have consistently demonstrated positive associations between midlife hypertension and dementia in later life. For instance, the Framingham Offspring Study^3^ and the Whitehall II^4^ cohort study found that elevated blood pressure in midlife, but not in later life, is linked to a higher incidence of dementia. However, clinical trials of antihypertensive medications have yielded inconclusive evidence regarding the causal link between hypertension and dementia. One study^5^ combing 14 RCTs (N=96,158) trialling antihypertensive medication interventions reported that lowering blood pressure was associated with a 7% reduced risk of dementia (OR= 0.93 [95%CI:0.88;0.98], *I*^2^ = 0.0%).

While, another study^6^ combing 9 RCTs (N=57,682) trialling a mix of antihypertensive and/or lifestyle interventions reported a similar 7% reduction in dementia risk (RR=0.93 [95%CI: 0.84;1.02], *I*^2^ =16%) but with confidence intervals that crossed the null, indicating uncertainty in the effect. Only three studies included in that review stratified their outcomes by dementia subtype. When pooled by subtype the effect sizes for AD and vascular dementia (VaD) were similar with confidence intervals that crossed the null which may reflect a lack of power in the stratified analyses. In addition, using large cohorts to emulate target trials in the investigation of antihypertensive medications and dementia risk has thus far yielded null^7^ or inconclusive^8^ results. Overall, these findings reflect conflicting evidence on whether lowering blood pressure reduces dementia risk and highlight the need for more studies stratifying outcomes by dementia subtype.

MR provides an alternative method to investigate the potential causal relationships between risk factors and disease outcomes by using genetic variants as instrumental variables for the risk factor. This approach helps to mitigate confounding factors and reverse causation that may introduce bias into observational studies. However, the evidence on blood pressure and dementia risk from MR studies has also been conflicting and inconclusive. Some studies have reported null associations ^9–11^, while others have reported a positive association between blood pressure and dementia risk, consistent with results from observational studies^12,13^. However, the majority of studies using an MR approach have reported an apparent protective direction of effect, with high blood pressure ostensibly reducing the risk of dementia^14–17^.

There are a number of potential explanations for these discrepant findings. One possible explanation is that the protective direction of effect is an artefact driven by survivor bias. One study^18^ reported a protective effect, albeit with confidence intervals crossing the null, of systolic blood pressure on AD and hypothesised that survivor bias may be an important factor accounting for their counter-intuitive results. That is, individuals who have hypertension are more likely to die prematurely from cardiovascular diseases before reaching an age where they may be vulnerable to receiving a diagnosis of AD. Supporting their hypothesis, in post-hoc analyses the study authors included genetic liability to coronary artery disease (CAD) in a multivariable MR model to account for a potential pleiotropic pathway via CAD and/or survivor bias. After conditioning on genetic liability for CAD the protective effect of genetically proxied high blood pressure attenuated towards the null.

Another plausible explanation is the use of different dementia outcomes across studies. Most observational studies use all-cause dementia as an outcome whereby many different subtypes of dementia are collapsed into one outcome category. By contrast MR studies thus far have mostly focused on the specific outcome of AD because these data are readily accessible in the form of publicly available genome wide association study (GWAS) summary data. It is likely that different subtypes of dementia which have vastly different underlying pathologies are differentially affected by various risk factors. For example, AD is primarily characterised by the accumulation of amyloid plaques and neurofibrillary tangles, leading to progressive neuronal damage and cognitive decline^19^. In contrast, VaD results from cerebrovascular damage, such as small vessel disease or stroke, which impairs blood flow to the brain and causes cognitive deficits^20^. Hypertension is an established causal risk factor for vascular damage, making it a probable direct causal risk factor for VaD^21^. However, its impact on AD may be less direct, potentially influencing the disease through vascular contributions to cognitive impairment or coexisting cerebrovascular disease. Therefore, hypertension may be a more important risk factor for VaD, directly contributing to brain atrophy and cognitive decline through its detrimental vascular effects.

In the current study we aimed to further investigate the relationship between high blood pressure and dementia. We employ observational analyses and individual-level MR analyses to investigate the relationships between hypertension and dementia, specifically focusing on all-cause dementia as well as the subtypes of AD and VaD, within the UK Biobank cohort. By leveraging genetic variants associated with blood pressure, we aim to clarify the role of systolic blood pressure (SBP) in the aetiology of these dementia subtypes. Specifically, we seek to address: is genetically proxied SBP associated with increased risk of all-cause dementia, AD and VaD?

## 2. Methods

Informed consent was provided by each participant on entry to the UK Biobank cohort. We followed the reporting guidelines provided by the Strengthening the Reporting of Observational Studies in Epidemiology - Mendelian Randomisation (STROBE-MR)^22^.

### 2.1 Participants

The participants came from the UK Biobank with over 500,000 participants. The participants, aged between 37 and 73 years at the time of recruitment, were enrolled between 2006 and 2010 across 22 assessment centres in the UK^23^. The baseline assessment gathered extensive data, including demographic, lifestyle, physical, biochemical and genotype data. In addition, the UK Biobank data are linked to the Hospital Episode Statistics (HES) dataset and death records^23^. Participants with a diagnosis of dementia from any cause at baseline were excluded from the present analyses.

### 2.2 Exposure Measures

Three different exposure measures were derived for analyses. These were: a continuous variable of mean SBP readings, and a binary variable of hypertension in the observational analyses, and a polygenic risk score (PRS) for blood pressure used as a genetic instrument in the one-sample MR analyses.

#### 2.2.1 Continuous blood pressure

A continuous blood pressure measure was derived by calculating the mean from two automatic SBP measurements taken at baseline. Where automatic measurements were missing, manual measurements were used. An adjusted SBP measure was created by adding 15 mmHg to the SBP and 10 mmHg to the diastolic blood pressure (DBP) for participants who were using anti-hypertensive medication at baseline, in line with previous research^24^.

#### 2.2.2 Binary hypertension

Participants were classified as having hypertension at baseline based on the following data at initial assessment: having a mean SBP reading at or above 140 mmHg or diastolic blood pressure reading at or above 90 mmHg^25^; HES data indicating diagnosis of hypertension; or self-reported use of anti-hypertensive medications (supplementary materials).

#### 2.2.3 Polygenic risk score for systolic blood pressure

A PRS for SBP was created as an instrumental variable (IV)^26^ and was derived based on a GWAS ^24^ identifying loci associated with SBP, which adjusted for age, sex and body mass index (BMI) as covariates. A total of 784 independent genetic instruments were selected after pruning based on a genome-wide significance threshold of *P*<5×10^−8^ and clumping, using the 1000 Genomes Project reference panel, retaining those with the lowest *P* value among those in linkage disequilibrium (*R*^2^=0.01). There was no overlap between the reference panel and the UK Biobank. Using the effect sizes provided by the initial GWAS by Evangelou et al.,^24^ and using a European reference panel, the following formula was used to calculate a weighted PRS for SBP for each individual *i*:

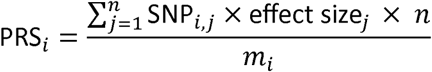

Where SNP*_i,j_* represents the genotype of SNP *j* (0, 1 or 2 effect alleles) for individual *i*; effect size*_j_* represents the effect size or beta regression coefficient of SNP *j* on SBP; *n* represents the total number of SNPs; and *m*_i_ represents the count of non-missing SNPs for individual *i*. Finally, we standardised the PRS by dividing by the standard deviation of SBP in the initial SBP GWAS such that all interpretation is with respect to one standard deviation change in the exposure for SBP.

#### 2.2.4 Covariates

The following covariates were included in the observational analyses: age, sex, smoking status (non-smoker, ex-smoker, light smoker, moderate smoker, and heavy smoker), and body mass index (BMI). Participants missing any study or covariate variable or those who responded with ‘Prefer not to answer’ for any variable were excluded from the analysis (Table 1).

**Table 1:** Characteristics of participants included in observational and MR analyses.

|  |  | Observational Analyses |  |  |  |  | MR Analyses |  |  |  |  |
| --- | --- | --- | --- | --- | --- | --- | --- | --- | --- | --- | --- |
|  |  | Total<br>(N=464,935) | Controls<br>(n=456,319) | All-Cause<br>Dementia<br>(n=8,616) | AD<br>(n=3,660) | VaD<br>(n=1,542) | Total<br>(N=447,915) | Controls<br>(n=439,660) | All-Cause<br>Dementia<br>(n=8,255) | AD<br>(n=3,512) | VaD<br>(n=1,470) |
| Sex, N female (%) |  | 254,410<br>(54.7) | 250,261<br>(54.8) | 4,149<br>(48.2) | 1,999<br>(54.6) | 638<br>(41.4) | 244,452<br>(54.6) | 240,479<br>(54.7) | 3,973<br>(48.1) | 1,919<br>(54.6) | 610<br>(41.5) |
| Age at recruitment, N (%) |  |  |  |  |  |  |  |  |  |  |  |
|  | <50 | 104,597<br>(22.5) | 104,496<br>(22.9) | 101<br>(1.2) | 32<br>(0.9) | 11<br>(0.7) | 99,942<br>(22.3) | 99,851<br>(22.7) | 91<br>(1.1) | 30<br>(0.9) | 10<br>(0.7) |
|  | 50-64 | 269,716<br>(58.0) | 266,350<br>(58.4) | 3,366<br>(39.1) | 1,428<br>(39.0) | 549<br>(35.6) | 260,278<br>(57.6) | 257,050<br>(58.5) | 3,228<br>(39.1) | 1,371<br>(39.0) | 520<br>(35.4) |
|  | >64 | 90,622<br>(19.5) | 85,473<br>(18.7) | 5,149<br>(59.8) | 2,200<br>(60.1) | 982<br>(63.7) | 87,695<br>(19.6) | 82,759<br>(18.8) | 4,936<br>(59.8) | 2,111<br>(60.1) | 940<br>(63.9) |
| Smoking status, N (%) |  |  |  |  |  |  |  |  |  |  |  |
|  | Non-smoker | 252,357<br>(54.3) | 248,356<br>(54.4) | 4,001<br>(46.4) | 1,799<br>(49.2) | 634<br>(41.1) | 243,185<br>(54.3) | 239,344<br>(54.4) | 3841<br>(46.5) | 1,731<br>(49.3) | 602<br>(41.1) |
|  | Ex-smoker | 166,276<br>(35.8) | 162,497<br>(35.6) | 3,779<br>(43.9) | 1,567<br>(42.8) | 720<br>(46.7) | 160,400<br>(35.8) | 156,783<br>(35.7) | 3,617<br>(43.8) | 1,731<br>(42.8) | 690<br>(46.9) |
|  | Light smoker | 12,652<br>(2.7) | 12,473<br>(2.7) | 179<br>(2.1) | 81<br>(2.2) | 32<br>(2.1) | 12,133<br>(2.7) | 11,961<br>(2.7) | 172<br>(2.1) | 76<br>(2.2) | 31<br>(2.1) |
|  | Moderate smoker | 14,668<br>(3.2) | 14,405<br>(3.2) | 263<br>(3.1) | 93<br>(2.5) | 68<br>(4.4) | 14,043<br>(3.1) | 13,794<br>(3.1) | 249<br>(3.0) | 88<br>(2.5) | 63<br>(4.3) |
|  | Heavy smoker | 18,982<br>(4.1) | 18,588<br>(4.1) | 394<br>(4.6) | 120<br>(3.3) | 88<br>(5.7) | 18,154<br>(4.1) | 17,778<br>(4.0) | 376<br>(4.5) | 113<br>(3.2) | 82<br>(5.6) |
| BMI (kg/m<sup>2</sup>), Mean (SD) |  | 27.4<br>(4.8) | 27.4<br>(4.8) | 27.8<br>(4.9) | 27.4<br>(4.7) | 28.8<br>(5.2) | 27.4<br>(4.7) | 27.4<br>(4.7) | 27.8<br>(4.8) | 27.4<br>(4.7) | 28.7<br>(5.1) |

|  | Observational Analyses |  |  |  |  | MR Analyses |  |  |  |  |
| --- | --- | --- | --- | --- | --- | --- | --- | --- | --- | --- |
|  | Total<br>(N=464,935) | Controls<br>(n=456,319) | All-Cause<br>Dementia<br>(n=8,616) | AD<br>(n=3,660) | VaD<br>(n=1,542) | Total<br>(N=447,915) | Controls<br>(n=439,660) | All-Cause<br>Dementia<br>(n=8,255) | AD<br>(n=3,512) | VaD<br>(n=1,470) |
| SBP (mmHg),<br>Mean (SD) | 138.0<br>(18.6) | 137.9<br>(18.6) | 144.1<br>(19.1) | 144.2<br>(18.7) | 145.0<br>(19.6) | 138.0<br>(18.6) | 137.9<br>(18.6) | 144.2<br>(19.2) | 144.3<br>(18.7) | 145.0<br>(19.6) |
| DBP (mmHg)<br>Mean (SD) | 82.2<br>(10.1) | 82.2<br>(10.1) | 81.7<br>(10.2) | 81.7<br>(10.0) | 82.1<br>(10.8) | 82.2<br>(10.1) | 82.2<br>(10.1) | 81.7<br>(10.2) | 81.7<br>(10.0) | 82.0<br>(10.7) |
| Antihypertensive<br>medication, N<br>(%) | 103,377<br>(22.2) | 99,692<br>(22.8) | 3,685<br>(42.8) | 1,374<br>(37.5) | 893<br>(57.9) | 99,944<br>(22.3) | 96,410<br>(21.9) | 3,534<br>(42.8) | 2,196<br>(37.5) | 851<br>(57.9) |
| Hypertensive<br>status, N (%) | 250,491<br>(53.9) | 244,046<br>(53.5) | 6445<br>(74.8) | 2,641<br>(72.2) | 1,287<br>(83.5) | 241,735<br>(54.0) | 235,556<br>(53.6) | 6,179<br>(74.9) | 2,539<br>(72.3) | 1,228<br>(83.5) |
| Missing, N (%) | 4,503 (1.0) | 4462 (1.0) | 41 (0.5) | 21 (0.6) | 8 (0.5) | 16 (0.0) | 16 (0.0) | 0 (0) | 0 (0) | 0 (0) |

AD: Alzheimer's disease; VaD: Vascular dementia; BMI: Body Mass Index; SD: Standard deviation; SBP: Systolic blood pressure; DBP: Diastolic blood pressure

### 2.3 Outcome Measures

Outcomes were classified as either; all-cause dementia; VaD or AD based on the International Classification of Diseases 9^th^ (ICD-9) or 10^th^ (ICD-10) revision codes provided via the linked HES data (Supplementary Table 1 and 2). Participants who received a diagnosis of both VaD and AD were excluded from the analyses where an exclusive diagnosis of either VaD or AD was the primary outcome. As elevated blood pressure is an established risk factor for increased risk of CAD, we used CAD as a positive control outcome.

### 2.4 Statistical Analyses

#### 2.4.1 Observational analyses

Logistic and Cox proportional hazard regression models were run to test the associations between three blood pressure traits (SBP, adjusted SBP and hypertension) and dementia outcomes (all-cause dementia, VaD and AD). Time to event was defined as the date of dementia diagnosis minus date of entry into the study. The models included covariates (age, sex, smoking status, and BMI) to adjust for potential confounding effects. Odds ratio (OR) and hazard ratio (HR) are reported with 95% confidence intervals (CI) per one standard deviation change in the exposure when considering SBP and adjusted SBP.

#### 2.4.2 MR analyses

A one-sample two-stage least squares individual-level MR analysis was performed to test the association between blood pressure and the three dementia outcomes (all-cause dementia, VaD and AD) using the PRS for SBP as the IV. In the first stage, we carried out a regression of SBP against the PRS in a linear model and in the second stage logistic regression and Cox regression models were used to test the association of the genetically predicted levels of SBP with the outcomes. Each stage was adjusted for age, sex and the first 20 principal components of genetic ancestry. To account for multiple testing a Bonferroni corrected *P* value threshold (.05/3 adjusting for three outcomes) was used to determine significance.

#### 2.4.3 MR assumptions

In any MR analysis, the validity of an instrumental variable is predicated upon three fundamental assumptions: (1) the genetic instrument must be associated with the risk factor (the relevance assumption), (2) the genetic instrument is not associated with any exposure and outcome confounder (the independence assumption), and (3) the genetic instrument must influence the outcome solely through the risk factor (the exclusion restriction assumption). With reference to the latter two assumptions, it is important to acknowledge that no reliable methods exist to confirm them directly; instead, only evidence refuting these assumptions can be sought. To evaluate the relevance assumption the associations between the PRS and the blood pressure traits were tested. A measure of instrument strength was calculated and an F-statistic value of >10 was considered to indicate that instruments are unlikely to be vulnerable to weak instrument bias^35^ To further test the relevance assumption, we evaluated the association of the PRS with CAD for which high blood pressure is one of the key established risk factors^36^. To mitigate violations of the independence assumption we restricted the analyses to European populations and adjusted for the first 20 principal components of genetic ancestry. To assess any violations of the exclusion restriction assumption we conducted pleiotropy-robust MR methods using the two-sample MR framework.

#### 2.4.4 Sensitivity analyses

Sensitivity analyses using pleiotropy-robust MR methods developed for summary-level data were conducted to assess the robustness of the potential causal estimates and detect potential biases. Summary statistics for all outcomes were generated from the output of the Cox regression models and merged and harmonized with genetic associations for SBP from the Evangelou et al. GWAS,^28^. The IV was created using the same 784 SNPs that were used to create the PRS for SBP in the main analyses. Two-sample MR analyses were then performed using the inverse-variance weighted (IVW), MR-Egger, Simple Median, and MR-PRESSO methods. In each case the resultant estimate represents the increase in HR per one standard deviation change in SBP.

In addition, we conducted a two-sample multivariable MR (MVMR) analysis^29^. In this analysis we used the IV for SBP, comprising of the initial set of 784 SNPs as used in the main analyses, and genetic associations with BMI. This additional analysis was carried out for two reasons. First, blood pressure and obesity are highly correlated traits and as such BMI is a potential pleiotropic pathway. The MVMR model allows us to attain an estimate for SBP after conditioning on potential pleiotropic pathways by including and thus accounting for genetic associations with BMI in the model. Second, most GWASs for blood pressure, including the Evangelou et al., GWAS^28^ that was used in current study, adjust for BMI in the creation of the GWAS. This adjustment has the potential to introduce bias in the MR estimate when both the exposure and outcome are influenced by the covariate^30^. However, MVMR models where the covariate is included as an additional exposure can provide an unbiased estimate of the direct effect ^30^. Therefore, we conducted the MVMR analysis including both SBP and BMI in the model to attain unbiased estimates of the association of genetic-proxied SBP and the three outcomes.

#### 2.4.5 Post-hoc analyses: Differential impact of VaD and AD

To further examine the potential differential impact of SBP on VaD and AD, we conducted two post-hoc analyses. First, Cochran’s Q statistic was calculated to assess heterogeneity between the effect sizes produced in the MR Cox analyses for VaD and AD. ^31^ Second, a logistic regression analysis was carried out with the PRS for SBP as the independent variable and a binary outcome (VaD vs AD) as the dependent variable, to assess if genetically proxied SBP can differentiate between those two dementia outcomes.

All analyses were performed using R version 2024.04.2+764 using the ivtools^32^, survival^33^ and the metafor^34^ packages. The analysis code is available at https://github.com/adawnir/BP-Dementia.

## 3 Results

### 3.1 Participant Characteristics

Data for 502,366 participants were extracted. Of these 204 participants had withdrawn consent and 83 participants had a diagnosis of dementia at baseline and were excluded from the present analyses. In addition, participants with missing data from any of the variables of interest were excluded from the analyses. Only participants who self-defined as White were included in the analyses. This left a total of 464,935 participants for inclusion in the observational analyses. After excluding participants with missing genetic data, a total of 447,915 were included in the MR analyses. See Table 1 for participant characteristics.

### 3.2 Observational analyses results

A summary of the observational analyses results can be found in Table 2. After applying a Bonferroni correction, SBP was not associated with any of the three dementia outcomes. However, once SBP was adjusted for use of antihypertensive medications, associations with increased risk were found for all three dementia outcomes. Similarly, the binary variable of hypertension was found to be associated with increased risk of all three dementia outcomes.

**Table 2:**
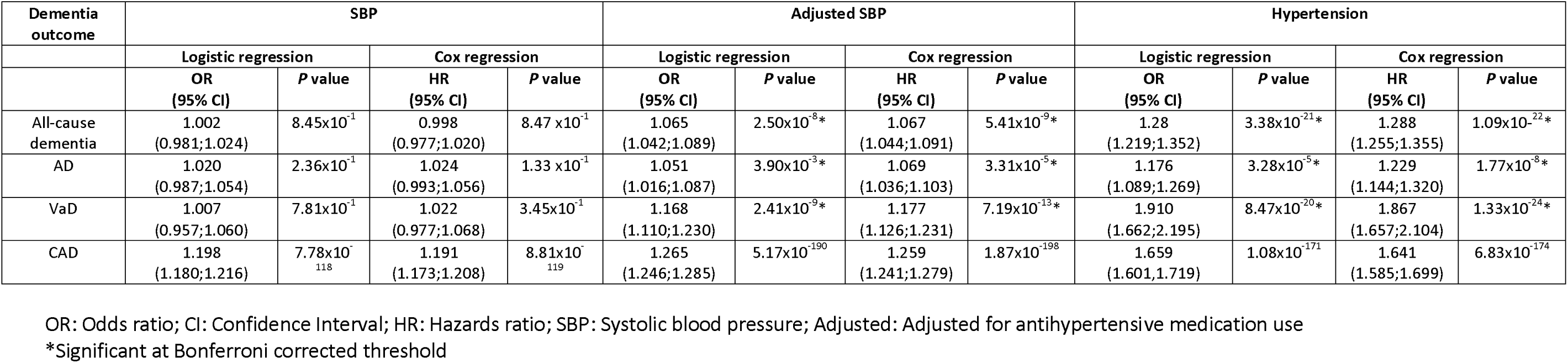
Results from observational analyses.

### 3.3 MR analyses results

The frequency distribution of individual PRS, for each outcome’s case and control groups, can be found in Figure 1.

**Figure 1:**
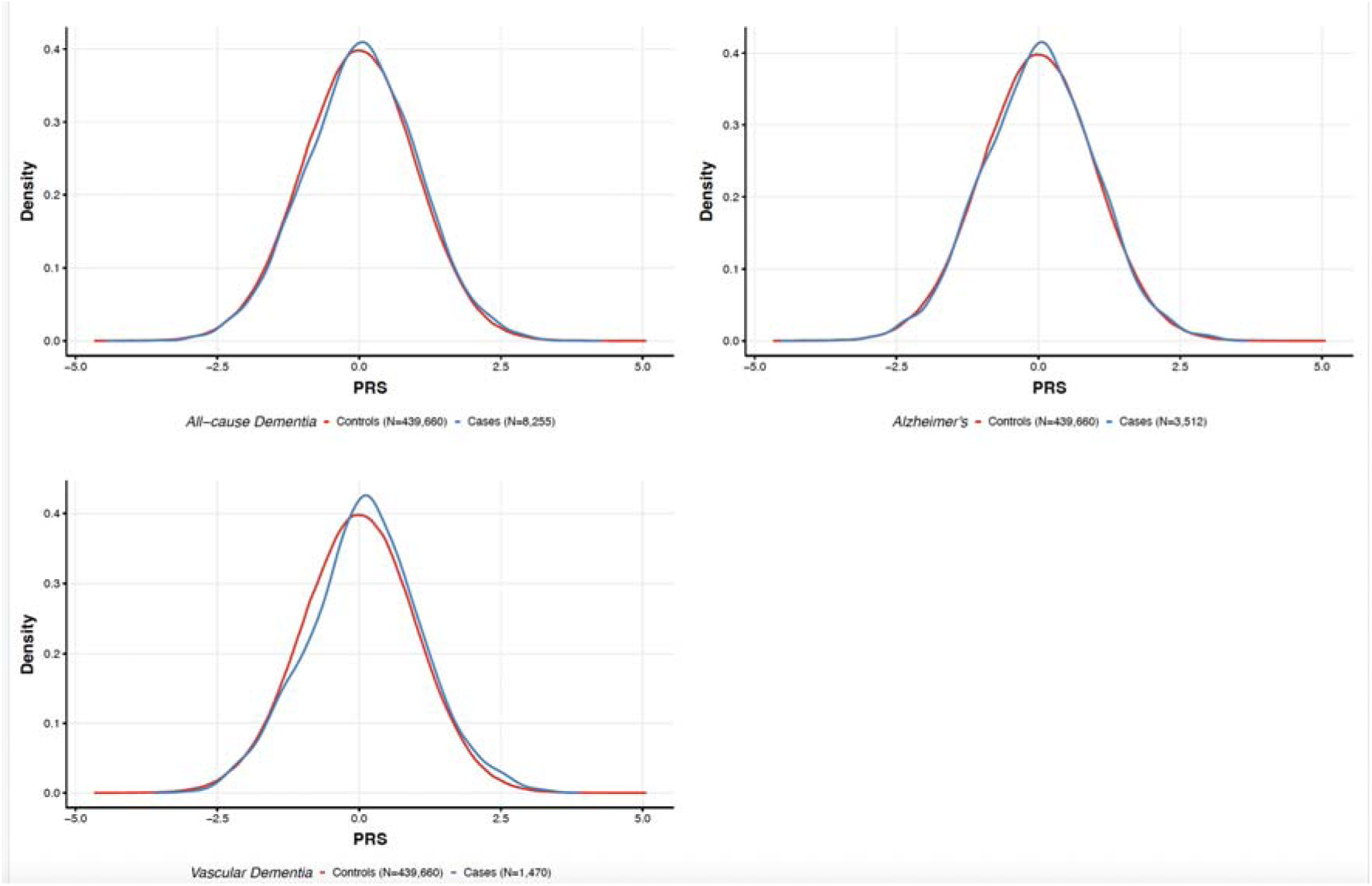
Polygenic risk score for systolic blood pressure (SBP) density plots distribution for controls and cases of all-cause dementia, VaD and AD

#### 3.3.1 MR assumption tests

The PRS was found to be significantly associated with all blood pressure traits. All F-statistic values were >10 indicating strong instrument strength (see Supplementary Table 3).

#### 3.3.2 MR two-stage least squares logistic and Cox regression results

For the outcome of all-cause dementia, we found an association between genetically-proxied SBP and increased risk of all-cause dementia in both the logistic and Cox models (OR 1.26 [1.15-1.38] and HR 1.28 [1.17-1.40]) (see Figure 2). However, when the two subtypes of interest were considered separately, there was found to be an increased risk association between genetically-proxied SBP and VaD in both the logistic and Cox models (OR=1.56 [1.25-1.93], HR=1.58 [1.28-1.97]). In contrast the 95% CI for AD included the null in both models (OR=1.10 [0.95-1.26], HR=1.11 [0.97-1.28]) (Table 3).

**Figure 2:**
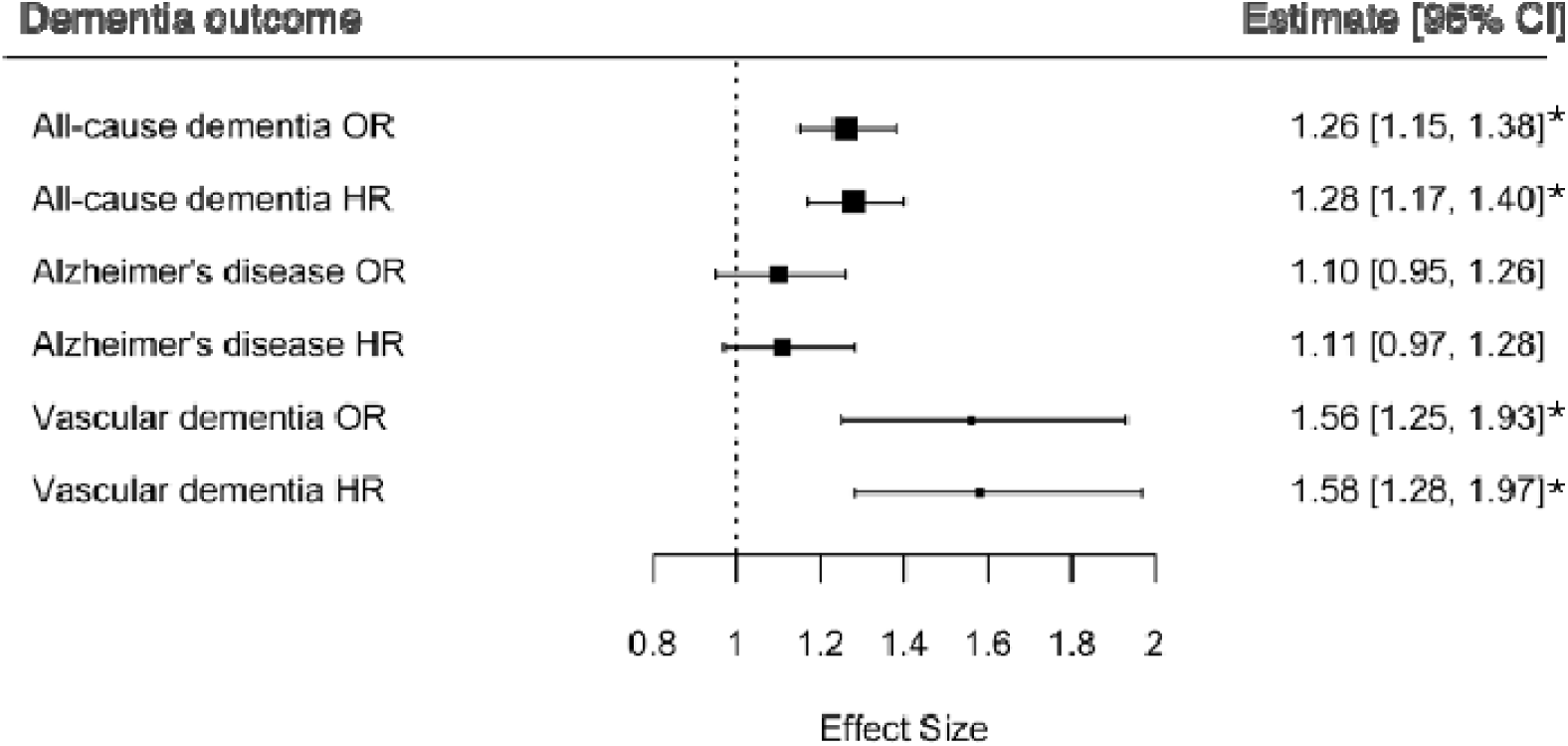
Forest plot for individual-level MR analyses for all-cause dementia, Alzheimer’s disease (AD) and vascular dementia (VaD), showing odds ratio (OR) for the logistic regression and hazard ratio (HR) for the Cox regression *Significant at Bonferroni corrected threshold

**Table 3:** Results from MR analyses.

| Dementia outcome | Logistic regression |  | Cox regression |  |
| --- | --- | --- | --- | --- |
|  | OR (95% CI) | P value | HR (95% CI) | P value |
| All-cause dementia | 1.26 (1.15;1.38) | $9.31 \times 10^{-7}$* | 1.28 (1.17;1.40) | $1.49 \times 10^{-7}$* |
| AD | 1.10 (0.95;1.26) | $1.91 \times 10^{-1}$ | 1.11 (0.97;1.28) | $1.24 \times 10^{-1}$ |
| VaD | 1.56 (1.25;1.93) | $6.21 \times 10^{-5}$* | 1.58 (1.28;1.97) | $2.97 \times 10^{-5}$* |
| CAD | 1.86 (1.74;1.98) | $4.03 \times 10^{-81}$* | 1.83 (1.72;1.95) | $1.27 \times 10^{-82}$* |

OR: Odds ratio; CI: Confidence Interval; HR: Hazards ratio; AD: Alzheimer's disease; VaD: Vascular dementia; CAD: Coronary artery disease
\*Significant at Bonferroni corrected threshold

#### 3.3.3 Sensitivity results

The results from the two-sample robust MR analyses (Supplementary Table 4) were consistent with the main analyses in the magnitude of the MR effect estimates for all-cause dementia (IVW HR=1.29, [95%CI: 1.15;1.44]) (see Figure 3). For VaD the IVW and MR PRESSO estimates were consistent with the main analyses (IVW HR=1.62 [95% CI:1.28;2.05]; MR-PRESSO HR=1.62 [1.28;2.05]). However, there was a small attenuation in effect with the simple median MR method (HR=1.40 [95% CI:0.99;1.98]). The MR-Egger intercept for all estimates yielded CIs which crossed the null indicating no evidence for unbalanced horizontal pleiotropy.

**Figure 3.**
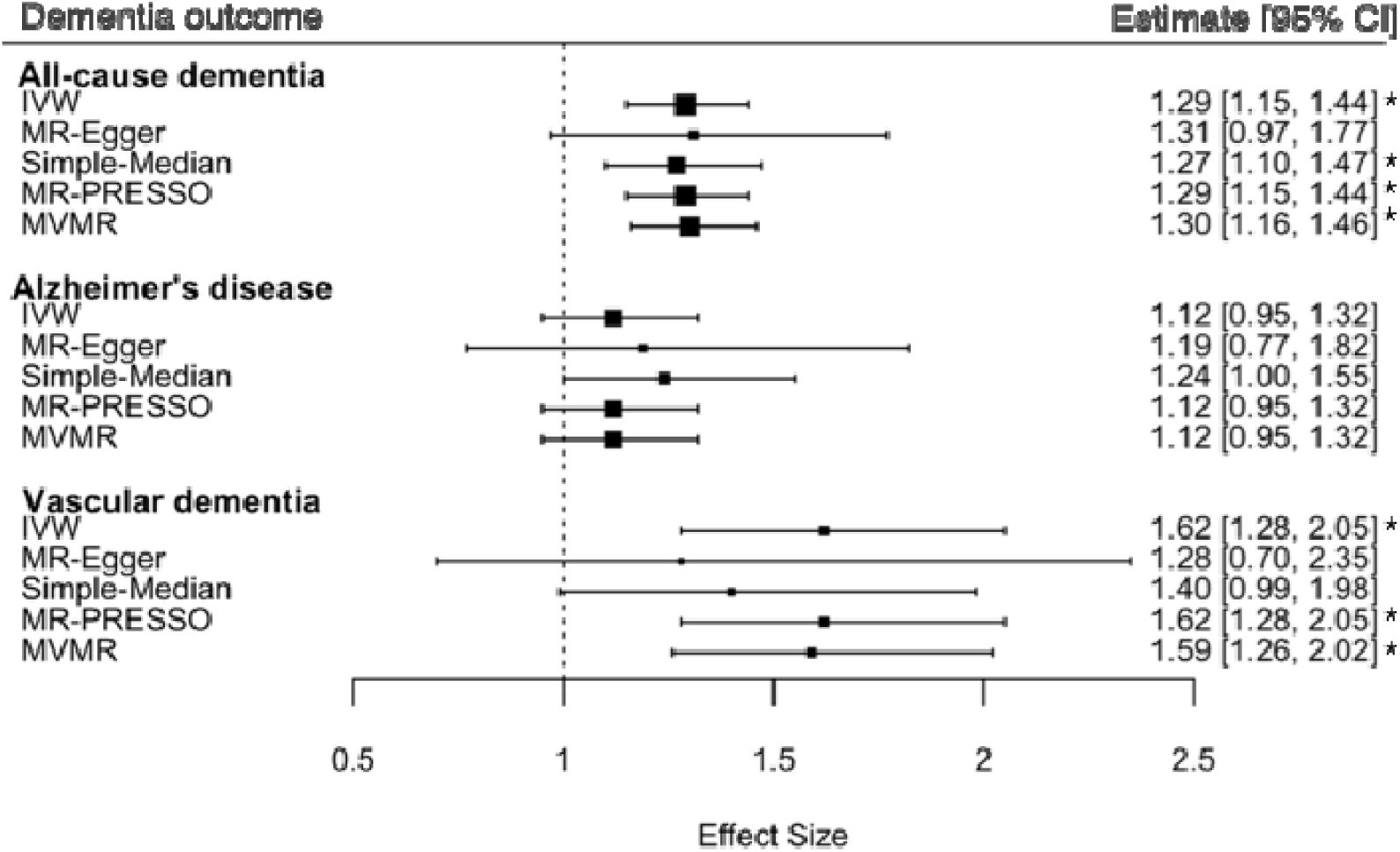
Forest plot of two-sample MR sensitivity analyses including univariable and multivariable MR models for all-cause dementia, Alzheimer’s disease (AD) and vascular dementia (VaD) *Significant at Bonferroni corrected threshold

In the MVMR models, the direct effect of SBP on all-cause dementia (HR=1.30 [95% CI: 1.16;1.46]) and VaD (HR=1.59 [95% CI:1.26;2.02]) after conditioning on genetic associations with BMI was similar in magnitude to the total effect observed in the univariable MR models (Supplementary Table 5).

#### 3.3.4 Positive control analysis using CAD

The association of the PRS and CAD was highly significant with similar magnitudes of effect in the two-stage least squares regression (HR= 1.83 [95% CI:1.72;1.95]) (Table 3) and the sensitivity analysis using robust summary-level MR approaches (IVW HR=1.90 [95% CI:1.74;2.08]) (Supplementary Table 4).

#### 3.3.5 Post-hoc results: Differential impact of VaD and AD

Cochran’s Q statistic indicated significant heterogeneity between the MR-derived Cox (X^2^=7.28, df=1, p=0.007) and logistic (X^2^=6.99, df=1, p=0.008) effect sizes for SBP on VaD and AD, tentatively indicating a differential impact of SBP on these subtypes. In a logistic regression model the PRS for SBP was associated with increased risk of VaD when compared to AD as the reference category (OR= 1.43; 95% CI: 1.11-1.83, p=0.006).

## 4 Discussion

We aimed to examine the relationship between raised blood pressure and dementia outcomes using observational and individual level MR analyses within the UK Biobank cohort. The observational analyses indicated that higher SBP, when adjusted for antihypertensive medication use, and the binary variable of hypertension were associated with an increased risk of all three dementia outcomes. The results from the MR analyses indicated that genetically-proxied SBP was associated with an increased risk of all-cause dementia. When the analyses were stratified into subtypes we observed an increased risk of VaD in the logistic and Cox models but not for AD. Post-hoc analyses provided support to the hypothesis that there is a differential effect of SBP on AD and VaD. This provides preliminary evidence that people with higher blood pressure are at increased risk of being diagnosed with VaD rather than AD. The positive associations between hypertension and VaD suggest that managing blood pressure may be particularly important for preventing VaD.

We conducted a number of post-hoc analyses which lend support to the hypothesis that there is potentially a differential effect of SBP on VaD vs AD. Specifically, there was significant heterogeneity between the effects for VaD and AD and that a higher genetic burden for increased SBP increases risk of VaD compared to AD by 43%. Although we cannot definitively conclude this from these results, it seems likely that the significant risk of genetically proxied SBP on all-cause dementia is driven via the effect of SBP on VaD. Our results support the hypothesis that the impact of genetically elevated blood pressure differs between dementia subtypes, with a more pronounced effect on the vascular pathology underlying VaD.

### Implications for further research

In light of the current findings, it is important to differentiate future studies examining dementia outcomes by subtypes rather than focusing on all-cause dementia. This recommendation is relevant to observational studies, RCTs and MR studies. Specifically, observational studies and RCTs should move away from collapsing dementia into one category as it is unclear if some studies reporting null^7^ associations between antihypertensive medications and all-cause dementia may have yielded different results if the outcomes were stratified by subtype. With regard to MR studies there is a pressing need for a GWAS for VaD to be created so the effects of genetically-proxied risk factors, especially those more pertinent to vascular mechanisms, can be investigated in relation to VaD risk.

This approach would ensure better understanding of risk factors, aid in the development of targeted interventions, and clarify the specific impact of raised blood pressure or other risk factors on each dementia subtype.

Furthermore, our individual-level data findings highlight a key limitation in using summary-level data for investigating age-related diseases. Specifically, many GWASs for AD e.g. Kunkle et al., (2019)^37^ restrict participants to those aged 65 years and over, potentially introducing selection bias when these data are merged with summary-level data on risk factors associated with earlier mortality.

### Clinical Implications

Antihypertensive treatments, may offer protective benefits against dementia, particularly VaD. It will be useful for further research to test this hypothesis in RCTs and MR studies using proxies for expression of drug target proteins. In addition to pharmacological interventions, promoting healthy behaviours and psychological well-being plays a crucial role in managing hypertension and reducing dementia risk^38^. Encouraging regular physical activity, a balanced diet^39^, stress reduction techniques, and adherence to treatment regimens may enhance the effectiveness of antihypertensive therapies. Moreover, psychological interventions aimed at improving health literacy and motivation may empower individuals to make sustained lifestyle changes^40^. Given the potential differential effects observed between VaD and AD, personalised approaches to dementia prevention may be indicated in the future. Tailoring hypertension management strategies to individual risk profiles and dementia subtypes could help to reduce an individual’s risk of dementia.

### Limitations

There are many strengths to the present study not least the very large sample size available in the UK Biobank. Despite this, numbers of rarer subtypes of dementia are still small and so it was not possible to extend the present analyses to other subtypes of dementia such as dementia with Lewy bodies or frontotemporal dementia. Another limitation is that the GWAS for SBP, which informed our PRS, was partially derived from UKB participants. Just under half (≈500,000) of the GWAS sample came from the UKB. Sample overlap can be a source of bias in MR studies; however, this bias can be mitigated with the use of a strong instrument^41^. The instrument strength in the current study was strong (*F*>1000) potentially limiting the bias from sample overlap. There is a potential for collider bias in the model as the genetic associations for SBP were adjusted for BMI in the GWAS. While this is a valid strategy to improve the detection of genetic signal for SBP this has been shown to potentially introduce collider bias in downstream causal inference^42^. This is especially so when the covariate potentially causes both the exposure and the outcome^43^. However, in the current study our further sensitivity analysis of conducting MVMR analyses ^30^ indicated little evidence of potential collider bias. A notable limitation to the UK Biobank is the sample population is not representative of the general population. Specifically, the cohort is comprised predominately of a highly educated, white sample who come from relatively high socio-economic backgrounds^44^. As the current study focused on analysing a sample of white participants the results may not generalise to other populations. This is an important point because some ethnic minority populations such as South Asians and Black communities are more vulnerable to risk factors such as hypertension^45^ therefore it is important to understand if this also puts them at a higher risk of VaD.

## Conclusion

In conclusion, our study provides preliminary evidence supporting a differential relationship between high blood pressure and different subtypes of dementia. Specifically, hypertension appears to be more strongly associated with increased risk of VaD. The differential effects observed between dementia subtypes highlights the need for future studies to move away from treating dementia as a unified outcome and to delineate subtypes into separate outcomes and to create a GWAS for VaD. Our work highlights the importance of understanding the potentially differential impact of risk factors on each subtype to ultimately develop personalised dementia risk reduction strategies for individuals.

## Author Contributions

VZ came up with the initial conceptualisation, the study design and analysis plan. JH, RW, MCH & VD were responsible for data curation. MA, MA, CC, SZ & FD conducted preliminary data analysis. RW, JH conducted the main analyses and RD validated the results. JH & RD were responsible for creating visualisations. RD was responsible for writing the manuscript with intellectual input and critical evaluation from all co-authors. VZ provided supervision.

## Data Availability

Data is available via the UK Biobank. Analytic code will be available via GitHub on publication of the manuscript in a peer reviewed journal.

## Acknowledgment

The present analyses were conducted under the UK Biobank Application No. 69328. Applications by researchers are made via application and approval is managed by the Data Access Management team. The work presented here relies on data generously provided by participants as well as that collected by the NHS as part of their care and support services.

## Supplementary Materials

### Supplementary Methods

**Supplementary Table 1.** Dementia diagnosis subtypes with corresponding ICD-9 codes.

| ICD-9 code | ICD-9 text | All-cause dementia | Alzheimer's dementia | Vascular dementia |
| --- | --- | --- | --- | --- |
| 290.2 | Senile dementia, depressed or paranoid type | ? |  |  |
| 290.3 | Senile dementia with acute confusional state | ? |  |  |
| 290.4 | Arteriosclerotic dementia | ? |  | ? |
| 291.2 | Other alcoholic dementia | ? |  |  |
| 294.1 | Dementia in other conditions classified elsewhere | ? |  |  |
| 331.0 | Alzheimer's disease | ? | ? |  |
| 331.1 | Pick's disease | ? |  |  |
| 331.2 | Senile degeneration of brain | ? |  |  |
| 331.5 | Creutzfeldt-Jakob disease | ? |  |  |

**Supplementary Table 2.** Dementia diagnosis subtypes with corresponding ICD-10 codes.

| ICD-10 code | ICD-10 text | All-cause dementia | Alzheimer's dementia | Vascular dementia |
| --- | --- | --- | --- | --- |
| A81.0 | Sporadic Creutzfeldt-Jakob disease | ? |  |  |
| F00 | Dementia in Alzheimer's disease | ? | ? |  |
| F00.0 | Dementia in Alzheimer's disease with early onset | ? | ? |  |
| F00.1 | Dementia in Alzheimer's disease with late onset | ? | ? |  |
| F00.2 | Dementia in Alzheimer's disease, atypical or mixed type | ? | ? |  |
| F00.9 | Dementia in Alzheimer's disease, unspecified | ? | ? |  |
| F01 | Vascular dementia | ? |  | ? |
| F01.0 | Vascular dementia of acute onset | ? |  | ? |
| F01.1 | Multi-infarct dementia | ? |  | ? |
| F01.2 | Subcortical vascular dementia | ? |  | ? |
| F01.3 | Mixed cortical and sub-cortical vascular dementia | ? |  | ? |
| F01.8 | Other vascular dementia | ? |  | ? |
| F01.9 | Vascular dementia, unspecified | ? |  | ? |
| F02 | Dementia in other diseases classified elsewhere | ? |  |  |
| F02.0 | Dementia in Picks disease | ? |  |  |
| F02.1 | Dementia in Creutzfeldt-Jacob disease | ? |  |  |
| F02.2 | Dementia in Huntington's disease | ? |  |  |
| F02.3 | Dementia in Parkinson's disease | ? |  |  |
| F02.4 | Dementia in HIV disease | ? |  |  |
| F02.8 | Dementia in other specified diseases classified elsewhere | ? |  |  |
| F03 | Unspecified dementia | ? |  |  |
| F10.6 | Mental and behavioural disorders due to use of alcohol - amnesic syndrome | ? |  |  |
| G30 | Alzheimer's disease | ? | ? |  |
| G30.0 | Alzheimer's disease with early onset | ? | ? |  |
| G30.1 | Alzheimer's disease with late onset | ? | ? |  |
| G30.8 | Other Alzheimer's disease | ? | ? |  |
| G30.9 | Alzheimer's disease unspecified | ? | ? |  |
| G31.0 | Circumscribed brain atrophy | ? |  |  |
| G31.1 | Senile degeneration of brain | ? |  |  |
| G31.8 | Other specified degenerative diseases of nervous system | ? |  |  |
| I67.3 | Binswanger's disease | ? |  | ? |

### List of antihypertensive medications

Amlodipine, Enalapril, Lisinopril, Losartan, Hydrochlorothiazide, Metoprolol, Valsartan, Diltiazem, Furosemide, Captopril, Atenolol, Carvedilol, Nifedipine, Spironolactone, Clonidine, Propranolol, Chlorthalidone, Verapamil, Hydralazine, Isosorbide mononitrate, Perindopril, and Ramipril.

## Supplementary Results

**Supplementary Table 3:** Relevance test: association between PRS and blood pressure variables using genetic instrument based on GWAS by Evangelou et al.,^24^.

| Variable | Beta<br>(95% CI) | P value | F-statistic | P value | R-squared |
| --- | --- | --- | --- | --- | --- |
| SBP | 15.53 (15.31;15.74) | $<2.23 \times 10^{-308} *$ | 1001.85 | $<2.23 \times 10^{-308} *$ | 0.045 |
| Adj SBP | 19.81 (19.57;20.05) | $<2.23 \times 10^{-308} *$ | 1328.41 | $<2.23 \times 10^{-308} *$ | 0.059 |

CI: Confidence Interval; SBP: Systolic blood pressure; DBP: Diastolic blood pressure; PP: Pulse pressure; Adj: Adjusted for antihypertensive medication use
\*Significant at Bonferroni corrected threshold

**Supplementary Table 4:** Results from the two-sample MR sensitivity analyses, where summary statistics from a time-to-event Cox proportional hazards model for each outcome were regressed onto summary statistics of genetic associations with systolic blood pressure (SBP) from Evangelou et al.,^24^ n=784 independent genetic variants associated with SBP were selected as instrumental variables. Effect sizes represent the total effect, that is the increase in hazard ratio (HR) when changing the exposure by one standard deviation unit.

| Outcome | Test | HR | 95% CI | P value |
| --- | --- | --- | --- | --- |
| All-cause dementia | IVW | 1.29 | 1.15;1.44 | $1.63 \times 10^{-05} *$ |
| | MR-Egger | 1.31 | 0.97;1.77 | $7.41 \times 10^{-2}$ |
| | Simple median | 1.27 | 1.10;1.47 | $1.08 \times 10^{-3} *$ |
| | MR-PRESSO | 1.29 | 1.15;1.44 | $1.83 \times 10^{-5} *$ |
| AD | IVW | 1.12 | 0.95;1.32 | $1.92 \times 10^{-1}$ |
| | MR-Egger | 1.19 | 0.77;1.82 | $4.37 \times 10^{-1}$ |
| | Simple median | 1.24 | 1.00;1.55 | $5.21 \times 10^{-1}$ |
| | MR-PRESSO | 1.12 | 0.95;1.32 | $1.09 \times 10^{-1}$ |
| VaD | IVW | 1.62 | 1.28;2.05 | $5.00 \times 10^{-5} *$ |
| | MR-Egger | 1.28 | 0.70;2.35 | $4.19 \times 10^{-1}$ |
| | Simple median | 1.40 | 0.99;1.98 | $5.58 \times 10^{-2}$ |
| | MR-PRESSO | 1.62 | 1.28;2.05 | $5.47 \times 10^{-5} *$ |
| CAD | IVW | 1.90 | 1.74;2.08 | $4.72 \times 10^{-45} *$ |
| | MR-Egger | 1.87 | 1.48;2.36 | $1.28 \times 10^{-7} *$ |
| | Simple median | 1.88 | 1.70;2.09 | $1.53 \times 10^{-33} *$ |
| | MR-PRESSO | 1.90 | 1.74;2.08 | $2.54 \times 10^{-40} *$ |

HR: Hazards Ratio; CI: Confidence Interval; AD: Alzheimer's disease; VaD: Vascular dementia; CAD: Coronary artery disease
\*Significant at Bonferroni corrected threshold

**Supplementary Table 5:** Results from Multivariable MR analyses, where summary statistics from a time-to-event Cox proportional hazards model for each outcome were regressed onto summary statistics of genetic associations with systolic blood pressure (SBP) and body mass index (BMI) from Evangelou et al.,^24^ and BMI summary statistics from Pulit et al.,^46^ n=784 independent genetic variants associated with SBP were selected as instrumental variables. Results represent the direct effect of SBP on the outcome after accounting for genetic associations with BMI. Effect sizes represent the increase in hazard ratio (HR) when changing the exposure by one standard deviation unit.

| Outcome | HR | 95%CI | P value |
| --- | --- | --- | --- |
| All-cause dementia | 1.30 | 1.16;1.46 | $1.03 \times 10^{-5}$* |
| AD | 1.12 | 0.95;1.32 | $1.93 \times 10^{-1}$ |
| VaD | 1.59 | 1.26;2.02 | $1.13 \times 10^{-4}$* |
| CAD | 1.92 | 1.80;2.10 | $8.74 \times 10^{-46}$* |

HR: Hazards Ratio; CI: Confidence Interval; AD: Alzheimer's disease; VaD: Vascular dementia; CAD: Coronary artery disease
\*Significant at Bonferroni corrected threshold

